# High stakes, low information, no roads: a qualitative evaluation of determinants of air ambulance utilization and decision making in rural Alaska

**DOI:** 10.64898/2026.09.21.26363602

**Authors:** Brian Rice, Brandon A. Berger, Robert Onders, Sar Medoff, Henry Saul

## Abstract

**Background:** In rural Alaska, where most communities lack road access, air ambulances serve as a critical link between remote communities and hospital services.

**Objective:** To examine how air ambulance utilization decisions are made within the rural Alaskan health organization ecosystem.

**Methods:** We conducted semi-structured interviews with five key stakeholders. Transcripts were analyzed thematically using the Practical, Robust Implementation and Sustainability Model (PRISM).

**Results:** Four themes emerged that collectively define the structure of the air ambulance decision-making system, each reflecting interactions across multiple PRISM domains: limited formal guidance, logistical constraints affecting transport options, variability driven by communication and clinician risk tolerance, and reliance on unwritten rules. Together, these describe decision-making in a limited information state with incomplete feedback, leading to experiential and locally evolved practices.

**Conclusion:** Air ambulance use reflects clinical–logistical constraints rather than critical illness alone. Linking transport decisions to outcomes and developing structured decision support may improve consistency and optimize care delivery.

## Introduction

Alaska is defined by its size and rurality. By far the largest state in the United States, its 570,641 square miles of land area are equivalent to either the area of California, Texas and Montana combined, or of France, Germany, Poland, and the United Kingdom combined. Given Alaska’s population is only about 738,000, this produces a population density (1.3 people per square mile) which is 970 times less than the highest density state (New Jersey) and 4.6 times less dense than the next lowest density state (Wyoming). While other highly rural U.S. states have remote communities (Census designated places) those communities are almost universally road accessible. Alaska is unique amongst all U.S. states in that 63.1% of its Census designated places have no road connection to the state highway system.^1^ This results in 20.8% (152,588 residents) of Alaska’s total 2020 population living in off-road communities.^2,3^ More than half (50.3%) of the individuals in off-road communities are American Indian and Alaska Native (AI/AN), compared to 14.3% in road-connected communities. Overall, nearly half (47.9%) of the state’s AI/AN population resides in off-road regions, with many living on traditional lands.

The lack of road access in these communities produces substantial barriers to emergency medical care access and results in excess mortality and specifically excess emergency care associated mortality in these regions.^4,5^ Finding ways for individuals living off the road to access timely medical care is perhaps the greatest clinical and logistical challenge facing the Rural Alaskan healthcare organizations (RAHOs) who operate the health facilities in these communities. Patient transport in these settings is not ancillary to care delivery but a core component.

To understand the nature of the access problem, the “spoke and hub” model of care in rural Alaska needs to first be understood. Many, but not all, RAHOs - which we define as health organizations operating in regions of the state that lack road connectivity to referral-level hospitals - serving these small communities are Tribal health organizations (THOs), which are managed by a local tribal entity, receive federal funding through the Indian Health Service, and work cooperatively with the Alaska Native Tribal Health Consortium (ANTHC) in Anchorage.

While services vary from one RAHO to another, they generally use a “spoke and hub” system.^6^ In this system, RAHOs run “spoke” clinics in smaller communities, administer a regional “hub” critical-access hospital (CAH) with physician staffing, laboratory, imaging, inpatient, and emergency department capabilities. As these CAH “regional hubs” typically lack surgical or specialty services, they depend on referrals and transfers to a tertiary “referral hub” hospital with subspecialty capacity in a major city such as Anchorage for definitive care in many cases.

Within this system structure, decisions about when and how to transfer patients between levels of care are continuous and unavoidable.

Care in “spoke” communities is most frequently delivered by Community Health Aides/Practitioners (CHA/Ps), a unique healthcare paradigm designed for rural Alaska in which community members train to serve as the “eyes and ears” of a clinician who manages the medical decision making for the patient remotely.^7^ In some cases, staffing is supplemented by nurse practitioners or physician assistants.^8^ Since “spoke” clinics have limited point-of-care testing, no radiology, no laboratory services, and a limited treatment options, the care team – a “hub” physician working in collaboration with the “spoke” clinician via phone and/or video conferencing – must determine whether the patients can be safely managed locally or require transport to a “hub”. These decisions are made under conditions of diagnostic uncertainty and constrained local resources.

“Spoke” communities are generally accessible year-round only by unpaved runway capable fixed-wing aircraft (with seasonal access by snow machine or boat). Patient movement therefore depends on either (a) commercial flights or (b) air ambulances. Commercial flights are infrequent, have limited seats, and lack medical staffing or monitoring. Air ambulances are a much more expensive and limited alternative resource which includes a team of pilots, flight nurses and medics, with advanced treatment options (including blood transfusions and life support) and monitoring. Determining how and when to use air ambulances creates exceptional challenges for RAHO clinicians providing emergency care. In the balance are clinical questions with incomplete information, limited resource allocation optimization concerns, and practical logistical considerations. This creates a low-feedback, high-uncertainty decision environment in which clinicians must continuously balance patient risk against system capacity.

What is known is that current patterns in these regions result in a per capita air ambulance utilization rate that approaches ground ambulance utilization rates in rural areas nationwide.^9^ This heavy use of air ambulances contrasts sharply with their role in the United States more generally, where aeromedical assets are generally reserved for rapid response to the highest acuity and most time-sensitive of emergencies.^10^ While there is no parallel to the Alaskan system within the US, there are other countries that have high reliance on air transport globally including Australia, Greenland, Finland, Norway, Sweden, and Canada.^11–14^ Limited work has been done to look at air ambulance decision-making in any of these countries to date.^15,16^ To our knowledge no work has yet been published centered on Alaskan air ambulance decision making.

The determinants of current air ambulance utilization practices remain poorly characterized, including the system-level barriers and facilitators that shape transport decision-making across clinical, logistical, and organizational contexts. Little is known about how clinicians make transport decisions in a setting defined by limited formal guidance, evolving informal norms, and incomplete outcome feedback. This reflects a broader gap in understanding how interacting system conditions influence decision-making in a low-feedback, high-uncertainty environment. The objective of this study was to characterize stakeholder perspectives on how current utilization practices have developed and to identify the factors that shape transport decision-making in this setting.

## Materials and methods

We conducted semi-structured 60 minute interviews with five key informants who represented critical stakeholders in the emergency medicine environment of rural Alaska: an experienced CHA/P trainer with more than 15 years of clinical experience, an experienced CHA/P clinic director with more than 15 years of clinical experience, a THO physician and rural service line director, a THO physician and chief medical officer, and the CEO of an air ambulance provider. Our interview guide was designed following the Practical, Robust Implementation and Sustainability Model (PRISM) framework to elucidate the contextual factors around the implementation of air ambulances from multiple perspectives. Interview questions also included direct questions about when air ambulances should and should not be used according to each interviewee’s perspective. Ethical approval was obtained from the Alaska Area Institutional Review Board (IRB), Stanford University IRB and Tribal approval was obtained from the Southcentral Foundation, the Alaska Native Tribal Health Consortium, and the Maniilaq Association.

## Results

Analysis of key informant interviews identified themes that were best understood through the interaction of four PRISM domains: (1) *intervention characteristics*, reflected in the largely undefined medevac decision process; (2) *external environment*, including roadless geography, weather, and transport availability; (3) r*ecipients*, including CHA/Ps, physicians, and medevac personnel with varying training and risk tolerance; and (4) *implementation and sustainability infrastructure*, including communication systems, the CHA/P care manual, and informal decision norms. These domains did not operate independently but interacted to shape air ambulance utilization decisions. Thematic analysis of our key informant interviews identified several themes about air ambulance decision making. Table 1 presents these themes mapped across PRISM domains, highlighting how multiple system components contribute to observed decision-making patterns.

**Table 1:** Thematic analysis.

| Theme | PRISM domains | Interpretation |
| --- | --- | --- |
| Limited formal guidance | Intervention Characteristics; Infrastructure | The decision process lacks formal specifications, tools, and training structures. |
| Air ambulance vs commercial decisions rely on experience/risk tolerance and logistics | External Environment; Recipients | Decisions require balancing patient acuity against aircraft availability, commercial schedules, monitoring needs, and clinician risk tolerance. |
| Communication challenges produce variability | Infrastructure; Recipients | CHA/P–physician communication is structurally asymmetric and dependent on individual skill. |
| Unwritten rules substitute for formal guidance | Infrastructure; Intervention Characteristics | Informal norms function as de facto decision support but are not standardized, auditable, or outcome-linked. |

These themes describe a real-world system in which limited formal guidance, external logistical constraints, individual clinician factors, and informal norms combine to produce highly variable air ambulance utilization decisions in a low-feedback, high-uncertainty environment. This variability arises not from isolated factors, but from the combined influence of system-level conditions lacking a standardized decision framework. Together, these findings indicate that air ambulance decision-making is an emergent property of interacting system constraints rather than a standardized clinical process.

**Table 2:**
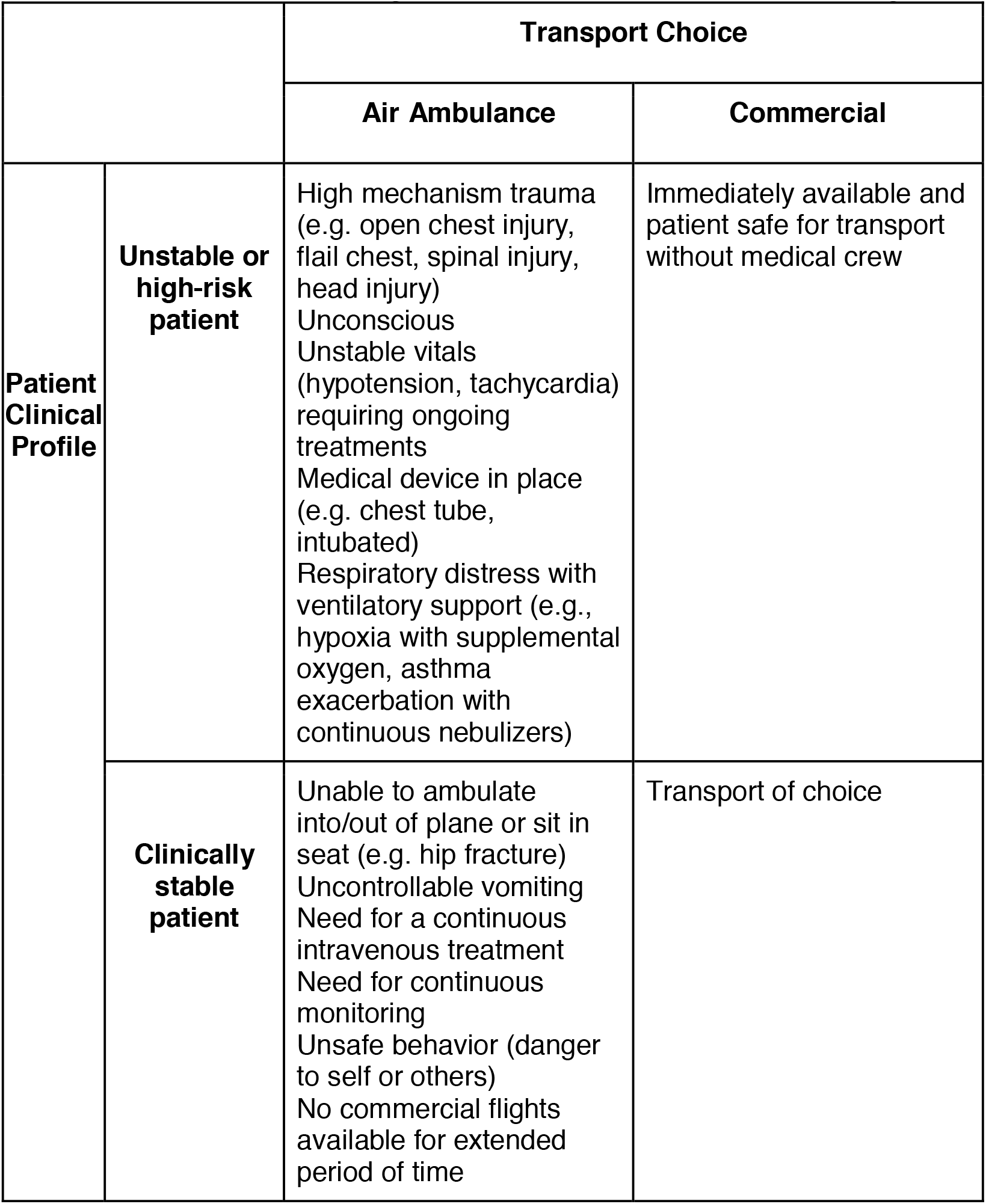
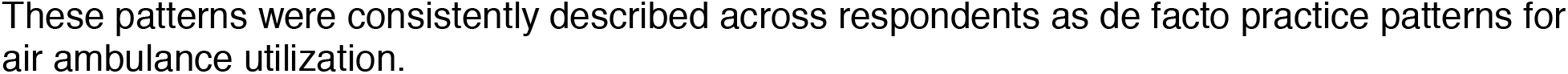
Informal commercial flight versus air ambulance decision making schema.

|  |  | Transport Choice |  |
| --- | --- | --- | --- |
|  |  | Air Ambulance | Commercial |
| **Patient Clinical Profile** | **Unstable or high-risk patient** | High mechanism trauma (e.g. open chest injury, flail chest, spinal injury, head injury)<br>Unconscious<br>Unstable vitals (hypotension, tachycardia) requiring ongoing treatments<br>Medical device in place (e.g. chest tube, intubated)<br>Respiratory distress with ventilatory support (e.g., hypoxia with supplemental oxygen, asthma exacerbation with continuous nebulizers) | Immediately available and patient safe for transport without medical crew |
|  | **Clinically stable patient** | Unable to ambulate into/out of plane or sit in seat (e.g. hip fracture)<br>Uncontrollable vomiting<br>Need for a continuous intravenous treatment<br>Need for continuous monitoring<br>Unsafe behavior (danger to self or others)<br>No commercial flights available for extended period of time | Transport of choice |

These patterns were consistently described across respondents as de facto practice patterns for air ambulance utilization.

### Air ambulance utilization decision-making is not formalized for physicians or non-physicians

Air ambulance utilization is not a structured component of clinical training for any part of the healthcare team, physician or non-physician. Existing emergency medical service decision systems (e.g. Medical Priority Dispatch System) are not designed for regions without roads. Air ambulance utilization off the road system is not the part of any medical school or residency curriculum. The guiding document for CHA/Ps clinical care is the Community Health Aid Manual (CHAM).^17^ While it mentions air ambulance considerations and some emergent situations when the CHA/P should “report NOW” to the “hub” to discuss transport, it does not house formal guidelines, algorithms, decision support, or pathways to provide a structured or generalized approach to making air ambulance utilization decisions. No similar manual exists for “hub” physicians receiving reports about patients possibly requiring transport. Physician decision-making regarding air ambulance use thus relies largely on individual experience and risk tolerance.

> *“I’m not aware of a good evidence-based guideline for helping someone to navigate [a medevac] decision. So, it’s oftentimes based on clinical experience. Which has all sorts of biases associated with it, as you know*.*” [Key Informant 2, THO Service Line Medical Director]*

### Decisions between air ambulance and commercial transportation are often challenging

The largest clinical challenge was commonly described for patients who are felt to require further evaluation or treatment available at the “hub” but are not obviously critically ill based on patient history, physical examination, and the limited point-of-care testing available in the “spoke” clinic. On the road system, these patients could be transported via private vehicle or ground ambulance for evaluation in an emergency department. However, without roads, commercial flights and air ambulances are the available options. Decisions between the two are complicated by a range of logistical realities and competing priorities. For example, commercial flights have limited seats and inflexible schedules, but in some instances may be a faster – though medically unmonitored – link to the emergency department based on air ambulance availability. Also, any use of an air ambulance represents an opportunity cost, as transportation of a more acutely ill or time sensitive patient may be delayed due to lack of local availability of air ambulances due to ongoing transport of another patient.

> *“If I can wait four hours and there’s no outpatient outcome difference for that patient, then what if I pull this aircraft out and then, all of a sudden, we have a stroke patient in our community where time really is of the essence?” [Key Informant 5, medevac provider CEO]*

### Communication challenges and individual risk tolerance produce high variability

The importance of communication was stressed by all five key informants. Organized and complete communication between “spoke” and “hub” is crucial for appropriate treatment and transport decisions about patients. However, this was identified as a challenge as the two halves of the team cooperating in a remote assessment are not only separate physically but often have very different training and levels of medical syntax. Individual communication skills play a role, but at a systems level, the CHAM provides structures to guide CHA/P reporting to “hub” clinicians but receiving clinicians do not have a reciprocal formalized structure for their communicating with the CHA/Ps. Together, these factors can result in variability in the fidelity of communication. The combination of a challenging communication environment with reliance on individual clinician judgment results in high inter-provider variability in air ambulance utilization decisions.

> *“The health aides are very structured in their approach through the Health Aide Manual. I think that we [physicians] are not very structured in our response*.*” [Key Informant, Chief medical officer, THO]*

### Unwritten rules exist about air ambulance utilization

Despite the absence of formal guidelines, participants described a shared set of informal decision patterns that have emerged based on accumulated clinical and logistical experience. Among these are scenarios that would not typically involve using an emergency aeromedical asset elsewhere but reflect the lack of other medical transport modalities available in rural Alaska.

These patterns were consistently described across respondents as de facto practice patterns for air ambulance utilization.

## Discussion

In rural Alaska, the absence of ground transport fundamentally alters the role of air ambulances. Air ambulance usage is not limited to critically ill patients meeting a narrow set of conditions but instead functions as a mechanism for resolving clinical–logistical mismatches, where patient needs cannot be safely accommodated by commercial transport or “spoke” clinic care.

Decisions in this space do not always adhere to conventional emergency medicine paradigms, and aspects of the clinical and logistical environment in which RAHOs operate create a large intermediate group of undifferentiated patients for whom transport decisions are uncertain.

Our findings suggest that this uncertainty arises from the interaction of four factors: a lack of formalized decision guidance, externally constrained transport options, variability of clinician experience and risk tolerance, and a limited information state at the time of evaluation.

In sum, these decisions exhibit characteristics of what Hogarth describes as “wicked” problem environments, or those characterized by incomplete information, delayed or absent feedback, and thus forcing decision-makers to act under uncertainty without clear or stable rules.^18^ At RAHOs, clinicians must make high-stakes transport decisions in low clinical information states, relying on variable communication fidelity, and with limited ability to observe outcomes, creating conditions where individual judgement, experiential learning, and locally evolved practices predominate.^19^

This study has important limitations. As a qualitative analysis of key informants, it does not link decision-making processes to patient outcomes and thus cannot determine the optimal balance between air ambulance and commercial transport. More broadly, air ambulance utilization decisions inherently require forecasting clinical trajectories under uncertainty, a constraint that cannot be fully eliminated. This work reflects the clinical realities and practice patterns of a single region of Alaska. Similar input from clinicians in other regions should inform the development and broader implementation of future protocols or processes.

Future work should focus on linking air ambulance utilization to downstream outcomes to establish an evidence base for transport decisions. This work also emphasizes a parallel opportunity to develop tools to standardize communication, structure risk assessment, and improve consistency in this unique rural emergency healthcare environment.

## Conclusion

Air ambulance utilization in rural Alaska reflects a complex interaction between clinical uncertainty, logistical constraints, and the absence of formal guidance. Addressing this variability will require both improved evidence which links transport decisions to outcomes and the development of structured approaches to support decision-making in resource-limited settings.

## Data Availability

All data produced in the present work are contained in the manuscript

